# Joint Detection of Serum IgM/IgG Antibody is An Important Key to Clinical Diagnosis of SARS-COV-2 Infection

**DOI:** 10.1101/2020.07.07.20146902

**Authors:** Fang Hu, Xiaoling Shang, Meizhou Chen, Changliang Zhang

## Abstract

**Background:** This study was aimed to investigate the application of SARS- COV-2 IgM and IgG antibodies in diagnosis of COVID-19 infection.

**Method:** This study enrolled a total of 178 patients at Huangshi Central Hospital from January to February, 2020. Among them, 68 patients were SARS-COV-2 infected confirmed with nucleic acid test (NAT) and CT imaging. 9 patients were in the suspected group (NAT negative) with fever and other respiratory symptoms. 101 patients were in the control group with other diseases and negative to SARS-COV-2 infection. After serum samples were collected, SARS-COV-2 IgG and IgM antibodies were tested by chemiluminescence immunoassay (CLIA) for all patients.

**Results:** The specificity of serum IgM and IgG antibodies to SARS-COV-2 were 99.01% (100/101) and 96.04% (97/101) respectively, and the sensitivity were 88.24% (60/68) and 97.06% (66/68) respectively. The combined detection rate of SARS-COV-2 IgM and IgG antibodies were 98.53% (67/68).

**Conclusion:** Combined detection of serum SARS-COV-2 IgM and IgG antibodies had better sensitivity compared with single IgM or IgG test, which can be used as an important diagnostic tool for SARS-COV-2 infection and a screening tool of potential SARS-COV-2 carriers in clinics, hospitals and accredited scientific laboratory.

## Introduction

The novel coronavirus (SARS-CoV-2) is a new virus responsible for an outbreak of respiratory illness known as COVID-19, and now it is a global pandemic in more than 215 countries [1]. The current standard diagnostic method for diagnosis of COVID-19 is to detect the virus nucleic acid RT-PCR [2]. However, real-time PCR detection had some limitations, e.g. time-consuming, complicated operation with specialized equipment, need special detection sites, which limit its application during COVID-19 outbreak [3].Therefore, a simple, sensitive and accurate test was needed to identify SARS-COV-2 infected patients in a COVID-19 outbreak area urgently.

Based on the diagnosis experience of many clinical cases, the detection of novel coronavirus antibody can be used as an auxiliary diagnosis of novel coronavirus pneumonia [4,5]. After SARS-COV-2 infection, the body’s immune system can create immune response to fight against the virus and produce specific antibodies. In general virology, the immunoglobulin M (IgM) antibody, produced in the early period after the infection, can indicate the current infection or the recent infection. Immunoglobulin G (IgG) antibody is also an important antibody produced by the immune system, indicating that the disease is in the middle to late stage, or presence of past infection. Therefore, the combined detection of IgM and IgG can be used not only in the early diagnosis of infectious diseases, but also in the assessment of the stage of infection.

Chemiluminescence immunoassay (CLIA) has been developed as an effective combination of immunoassay and chemiluminescence system [6] and it has been used recently in SARS-COV-2 diagnosis. In this study, we used a CLIA test product, which can detect IgM and IgG in human serum within 30 minutes. Our aim is to investigate the clinical value of CLIA for the diagnosis of SARS-COV-2 infection. This CLIA method demonstrated good sensitivity and specificity in our study, which can be used not only in hospitals and accredited laboratories, but also can be used in airports, border ports, seaports, and train stations etc. This CLIA method has potential to be a powerful weapon against the COVID-19 pandemic.

## Materials and Methods

### Patients

This study enrolled a total of 178 patients who visited Huangshi Central Hospital in Hubei Province, China, between January and February 2020. The patients included 91 males (51.1%) and 87 females (48.9%) with a mean age of 54.3 years (ranging from 2 months to 94 years). Among them, SARS-COV-2 group had 68 patients, 36 males and 32 females (ranging from 30 years to 90 years); suspected group had 9 patients, 7 males and 2 females (ranging from 2 months to 64 years); negative group had 101 patients, 48 males and 53 females (ranging from 2 years to 94 years). This study is in compliance of ICC clinical trial specifications and the Helsinki Declaration.

### Serologic tests

Serum was collected from all patients. Serum SARS-CoV-2 IgG and IgM were tested by CLIA kits and iFlash 3000 fully automated CLIA analyzer from Shenzhen YHLO Biotech Co., Ltd (China). Briefly, serum was separated by centrifugation at 2500g for 5 min within 12 hours of collection. The magnetic beads of these CLIA assays are coated with two antigens of SARS-CoV-2 (Nucleocapsid protein and N protein, Spike protein and S protein). SARS-CoV-2 IgM/IgG titers (in arbitrary units, AU/ml) were calculated automatically by the CLIA analyzer based on relative light units (RLU), and the viral antibody titer is positively associated with RLU. The cut-off values for positive SARSCoV-2 IgM and IgG are both 10 AU/ml.

### SARS-COV-2 nucleic acid test

RT-PCR was used to detect open reading frame 1ab (ORF1ab) and nucleocapsid protein (N) in the SARS-COV-2 genome. CT value interpretation of test results is based on instruction of use from manufacturer. Confirmation of positive COVID-19 is based on at least one target-specific RT-PCR positive result of ORF1ab and N genes of SARS-COV-2 in the same specimen.

### Data analysis

The statistical analysis was performed using SPSS 19.0 statistical software (IBM SPSS, Chicago, IL, USA). The kappa coefficient was calculated. Kappa ≥0.75 indicates good consistency, 0.75≥ kappa >0.4 for medium consistency, and kappa <0.4 for poor consistency.

The specificity and sensitivity of the CLIA test kits were calculated according to the following equations:

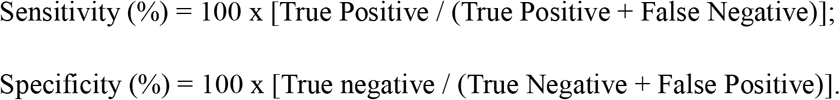

## Results

### The specificity of SARS-CoV-2 IgG/IgM antibody test

Samples from both NAT negative patients (suspect group, 9 subjects) and other diseases population (control group, 101 subjects) were used to assess the clinical specificity of the assay (Table 1). Among the 101 patients in the control group, 100 patients were negative for SARS-COV-2 IgM antibody, with a clinical specificity of 99.01% (100/101). 97 patients were negative for SARS-COV-2 IgG antibody, with a clinical specificity was 96.04% (97/101). In the suspect group, all 9 patients had negative antibody test results. The false positive results of SARS-COV-2 IgM and IgG antibodies may be caused by autoantibodies, heterophilic antibodies and other factors.

**Table 1:** Clinical specificity of SARS-CoV-2 IgM and SARS-CoV-2 IgG

| Sample | SARS-COV-2 IgM | SARS-COV-2 IgG |
| --- | --- | --- |

|  | Number | Negative | Positive | Clinical<br>Specificity | Negative | Positive | Clinical<br>Specificity |
| --- | --- | --- | --- | --- | --- | --- | --- |
| Suspect<br>Group | 9 | 9 | 0 | 100.00% | 9 | 0 | 100.00% |
| Control<br>Group | 101 | 100 | 1 | 99.01% | 97 | 4 | 96.04% |

### The detection sensitivity of SARS-CoV-2 IgG/IgM antibody test

Samples from 68 SARS-Cov-2 infected patients (confirmed with RT-PCR) were used to evaluate the clinical sensitivity of the assays (Table 2). We analyzed the clinical sensitivity on both SARS-CoV-2 IgM and IgG antibodies at three time periods, before 7 days, 7-14 days, and after 14 days since the onset of the symptoms. During these time periods, SARS-CoV-2 IgM demonstrated clinical sensitivity of 75.00%, 88.00%, and 93.55% respectively, and SARS-CoV-2 IgG demonstrated 83.33%, 100.00%, and 100.00% respectively. The total clinical sensitivity of SARS-COV-2 IgM and IgG to SARS-COV-2 infection were 88.24% (60/68) and 97.06% (66/68), respectively.

**Table 2:**
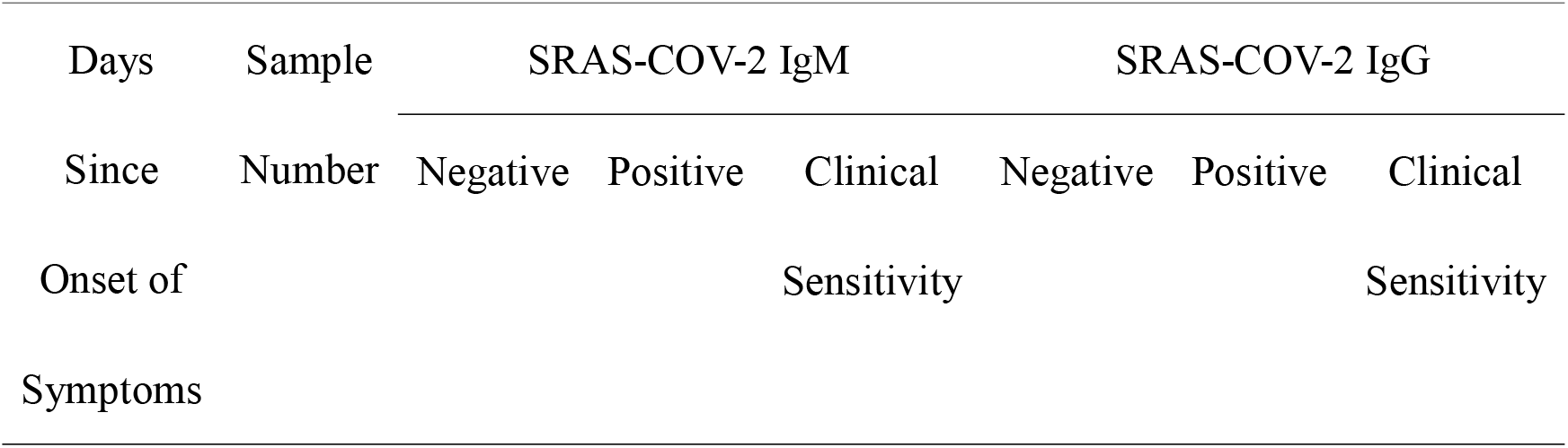
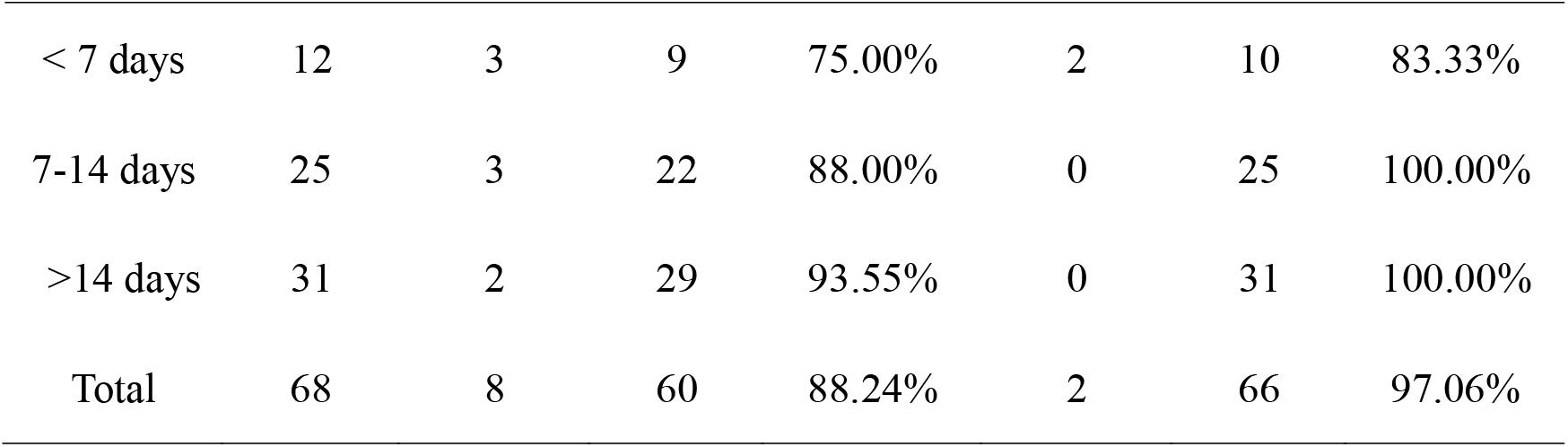
Clinical sensitivity of SARS-CoV-2 IgM and SARS-CoV-2 IgG

| Days | Sample | SRAS-COV-2 IgM |  |  | SRAS-COV-2 IgG |  |  |
| --- | --- | --- | --- | --- | --- | --- | --- |
| Since | Number | Negative | Positive | Clinical | Negative | Positive | Clinical |
| Onset of |  |  |  | Sensitivity |  |  | Sensitivity |
| Symptoms |  |  |  |  |  |  |  |
| < 7 days | 12 | 3 | 9 | 75.00% | 2 | 10 | 83.33% |
| 7-14 days | 25 | 3 | 22 | 88.00% | 0 | 25 | 100.00% |
| >14 days | 31 | 2 | 29 | 93.55% | 0 | 31 | 100.00% |
| Total | 68 | 8 | 60 | 88.24% | 2 | 66 | 97.06% |

### Comparison of SARS-COV-2 antibody test and SARS-COV-2 nucleic acid test (NAT)

The comparison between SARS-COV-2 IgM / IgG antibody test and NAT of 178 patients is shown in Table 3. The positive predictive value of SARS-COV-2 IgM / IgG antibody detection was 93.06% (67/72), the negative predictive value was 99.06% (105/106). The positive predictive value of the NAT for SARS-COV-2 was 100% (68/68) and the negative predictive value was 96.36% (106/110).

**Table 3:**
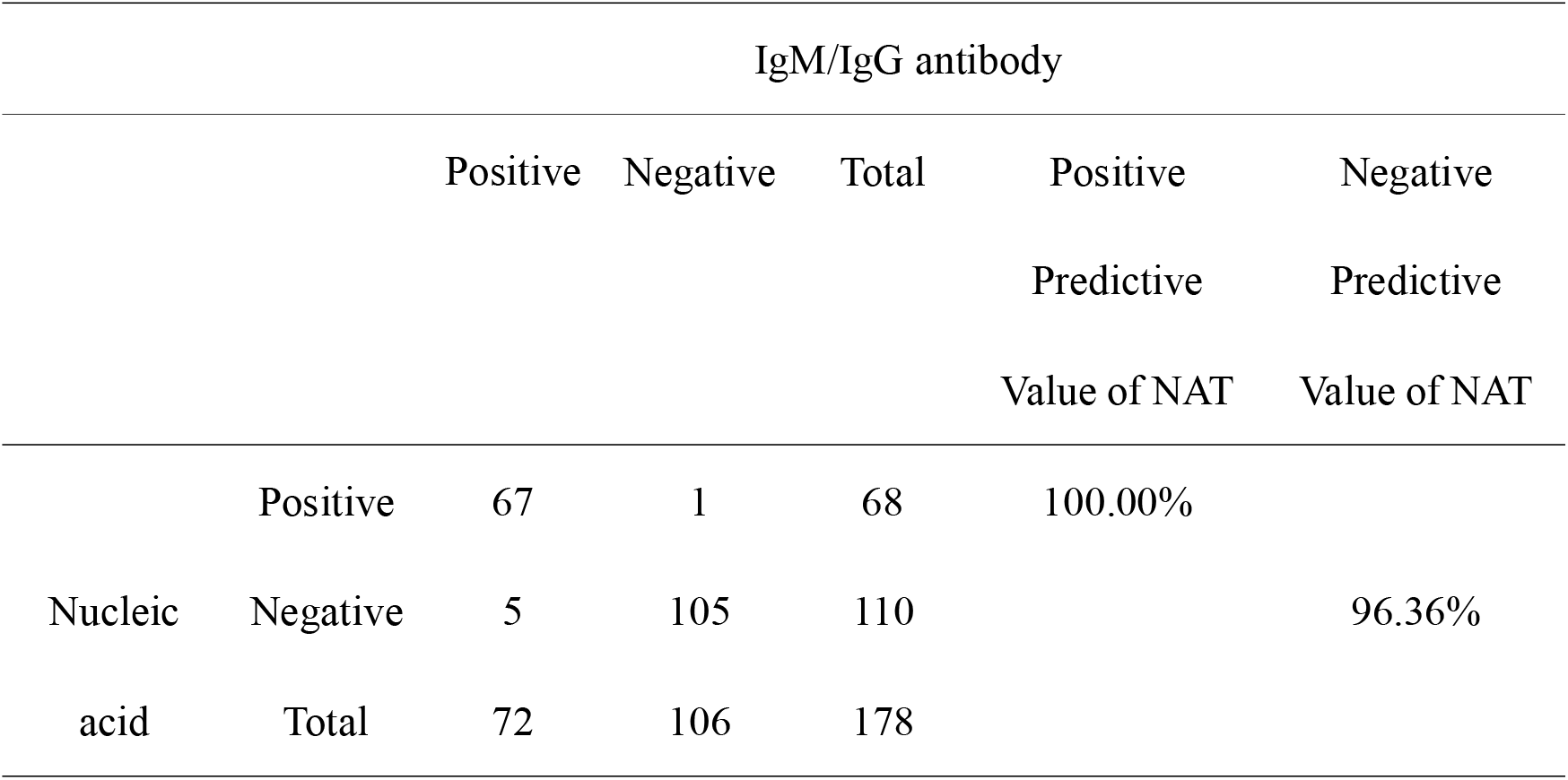
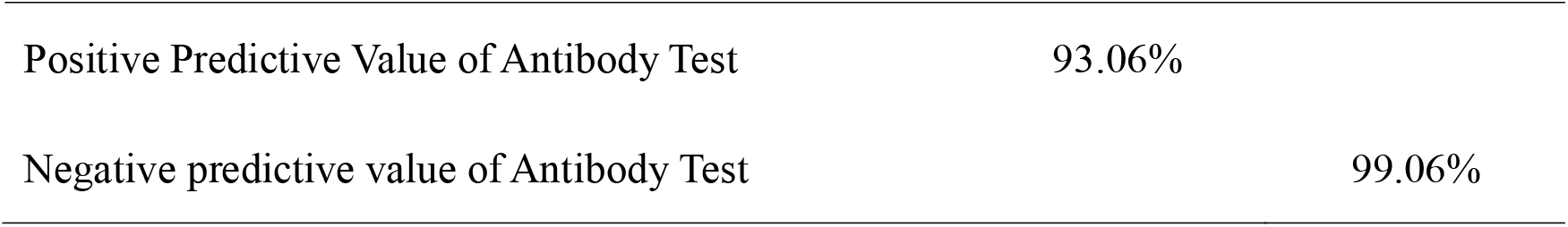
Comparison of SARS-COV-2 IgM/IgG antibody detection and SARS-COV-2 nucleic acid detection

|  |  | IgM/IgG antibody |  |  |  |  |
| --- | --- | --- | --- | --- | --- | --- |
|  |  | Positive | Negative | Total | Positive Predictive Value of NAT | Negative Predictive Value of NAT |
| Nucleic acid | Positive | 67 | 1 | 68 | 100.00% |  |
|  | Negative | 5 | 105 | 110 |  | 96.36% |
|  | Total | 72 | 106 | 178 |  |  |

|  |  |
| --- | --- |
| Positive Predictive Value of Antibody Test | 93.06% |
| Negative predictive value of Antibody Test | 99.06% |

## Discussion

Novel Coronavrius is caused by severe acute respiratory syndrome coronavirus 2 (SARS-CoV-2) [7]. SARS-COV-2 belongs to the subfamily of *Coronavirinae* (named *Betacoronavirus)*, and its genome is a single-stranded positive-sense RNA [8]. Different from MERS-CoV and SARS-CoV, SARS-COV-2 is the seventh member of the family of coronaviruses that infect humans [9]. The disease (COVID-19) is spreading rapidly within five months, and now it has been found in more than 215 countries. By June 19th, 2020, over 8, 300, 000 COVID-19 confirmed patients were reported, with□>□450, 000 deaths. Currently, the SARS-COV-2 NAT is the routine confirmation test for the clinical diagnosis of COVID-19 [2]. However, not all clinical COVID-19 patients might have positive result from SARS-COV-2 NAT. The reasons of false negative NAT results in COVID-19 include collection and storage of the sample, the condition of NAT test laboratory and the quality of the test kit [10]. Therefore, nucleic acid detection, CT imaging, blood routine examination and other methods can be used together for the comprehensive diagnosis of COVID-19.

Since February 2020, several SARS-COV-2 IgM and IgG antibody immunoassay kits have been developed in China. Antibody detection is a new detection method for SARS-COV-2, so the clinical specificity and clinical sensitivity of such tests must be carefully validated [11]. Our study demonstrated that the SARS-COV-2 IgM and IgG CLIA kits (YHLO Biotech, Shenzhen, China) had a high clinical specificity, reaching 99.01% and 96.04% respectively. Therefore, SARS-COV-2 IgM and IgG antibody detection reagents have high clinical specificity and can meet the screening and diagnosis requirements of SARS-COV-2.

In COVID-19 cases, the clinical sensitivity of SARS-COV-2 IgM detection was 88.23%, while the clinical sensitivity of SARS-COV-2 IgG detection was 97.06%. CLIA system can simultaneously detect 150-300 clinical samples, which is a good tool for screening and diagnosis of the Novel Coronavirus Pneumonia infected by SARS-COV-2. Our study results showed that the combined detection of SARS-COV-2 IgM and IgG antibodies is an effective tool to improve the diagnostic sensitivity and specificity and reduce the chance of false negative NAT results. We demonstrate that the antibody detection can be used as one of the effective methods of COVID-19 clinical detection.

In NAT confirmed group, serum from 68 COVID-19 cases were tested by SARS-CoV-2 IgM and IgG. SARS-CoV-2 IgM antibodies can be detected in 75.00% of patients before 7 days since onset of the symptoms, and the positive rate reached to the 88.00% on the period of 7-14 days, and then increased to 93.55% after 14 days. The positive rate of SARS-CoV-2 IgG was 83.33% before 7 days since onset of symptoms, and reached to 100.00% on 7-14 days and remained 100% after 14 days. In general, the immune response to infection by pathogenic microorganisms is first expressed as an increase in IgM antibody titer and then a rapid decrease until it disappears. While the IgG antibody titer normally increased in the middle and late stages of the infection, and it can be positive for a long time even after recovery. According to the results of this study, the positive rate of IgM in SARS-COV-2 infected patients is lower than IgG, because most of the infected patients were in the middle stage of infection or in the recovery stage. Interestingly, we have observed a phenomenon that SARS-CoV-2 IgM and IgG antibodies developed almost simultaneously and this observation is consistent with some recent studies [12]. Further studies are needed to verify this phenomenon in the diagnosis and prognosis of COVID-19.

We found false negative results for IgM / IgG in the NAT group. There might be three reasons: First of all, false negative results may be due to low antibody titer. When IgM and IgG titer is below the detection limit, the test result might be negative. Secondly, the difference in individual immune response and antibody production could be another reason for the false negative results in COVID-19 patients. The last reason might be that IgM antibody might decrease or even, disappear after 15 days. In each individual case, it is difficult to know exactly when or how long the patient has been really infected, and someone might have IgM titer below detection limit and not detectable. In the joint detection of SARS-COV-2 IgM and IgG, there was only one negative patients (male, 77 years old) who had respiratory failure, chronic obstructive pulmonary disease, coronary atherosclerosis, acute myocardial infarction, heart failure with SARS-COV-2 infection. In the control group, 5 cases were positive for antibody detection (1 case for IgM, 4 cases for IgG). The results suggested that the patients who had some other diseases, including tumors, leukemia, diabetes, hypertension, coronary atherosclerosis, bronchitis or lung infections might be more susceptible to infected by SARS-COV-2 and led to a positive antibody detection. Also, there might be false negative nucleic acids, or recovered/ mild / asymptomatic patients with SARS-COV-2. All of these cases will provide valuable reference for the follow-up study and clinical diagnosis of COVID-19.

Our study also has some limitations. For example, we didn’t investigate the cross-reaction with other pathogens (e.g., hCoV-NL-63 or others), MERS-COV, SARS-COV and some auto-antibodies that could cause interference for immunoassay. We also didn’t perform dynamic monitoring of the change of antibodies titer for in-depth study.

## Conclusions

Overall, testing SARS-CoV-2 IgG and IgM by CLIA method is convenient for sampling, and it has high efficiency. The results of this study indicated that combined detection of serum IgM and IgG antibodies to SARS-COV-2 had better sensitivity and specificity compared with single IgM or IgG test. Therefore, the serological test results can be used as an effective diagnostic tool for SARS-COV-2 infection. It also can be used as an efficient supplement of RNA detection for confirmation of SARS-CoV-2 infection in clinics, hospitals and accredited scientific laboratory.

## Data Availability

The datasets used and/or analyzed during the current study are available from the corresponding author on reasonable request.

## Abbreviations

SARS-CoV-2: severe acute respiratory syndrome coronavirus 2
NAT: nucleic acid test
CLIA: chemiluminescence immunoassay
IgM: Immunoglobulin M
IgG: Immunoglobulin G

## Acknowledgements

All the SARS-COV-2 IgG and IgM CLIA kits used in this study were kindly supplied by the manufacturers, namely, YHLO Biotech (Shenzhen, China).

## Disclaimers

The authors confirm that there are no conflicts of interest.

## Authors’ contribution

Fang Hu analyzed the data, drafted the article and contributed to study design. Xiaoling Shang and Changliang Zhang contributed to data gathering. Meizhou Chen contributed to study design, editing and revising the paper. All authors read and approved the final manuscript.

## Consent for publication

Not applicable.

